# Exposure to bovine livestock and latent tuberculosis infection in children: investigating the zoonotic tuberculosis potential in a large urban and peri-urban area of Cameroon

**DOI:** 10.1101/2024.02.06.24302437

**Authors:** Martine Augusta Flore Tsasse, Henry Dilonga Meriki, Hugues Clotaire Nana Djeunga, Marius Ambe Ngwa, Henri Olivier Tatsilong Pambou, Raïssa Dongmo, Ouethy Nguessi, Joseph Kamgno, Jane Françis Tatah Kihla Akoachere, Patrick Nguipdop Djomo

**Affiliations:** Department of Microbiology and Parasitology, Faculty of Science, University of Buea, Buea, Cameroon; Higher Institute for Scientific and Medical Research (ISM), Yaoundé, Cameroon; Ministry of Public Health, Jamot Hospital, Yaoundé, Cameroon; Department of Public Health, Faculty of Medicine and Biomedical Sciences, University of Yaoundé I, Yaoundé, Cameroon; Faculty of Epidemiology and Population Health, London School of Tropical Medicine and Hygiene, London, United Kingdom

**Author notes:** Corresponding author (PND).

**Keywords:** bovine tuberculosis, neglected zoonotic diseases, Latent tuberculosis Infection, children, *Mycobacterium bovis*, Interferon Gamma Release Assays, Cameroon, Africa

## Abstract

**Background:** Bovine tuberculosis (bTB), a neglected zoonotic disease, is endemic in cattle in many Sub-saharan African countries, yet its contribution to tuberculosis (TB) burden is understudied. Rapid urbanisation and increase in demand for animal proteins, including dairy products, increases the risk of spill over. This study compared the latent tuberculosis infection (LTBI) risk in children, a proxy-measure for recent TB infection, in children living in high cattle density areas to children from the general population in Cameroon.

**Method:** Cross-sectional study in the Centre Region of Cameroon in 2021, recruiting 160 children aged 2-15 years, stratified by exposure to livestock, people treated for pulmonary TB (PTB) and the general community. Veinous blood was tested for LTBI using QuantiFERON–TB Gold-Plus. Prevalences were calculated and the association to exposure and other risk factors investigated using logistic regression models.

**Results:** The crude LTBI prevalence were 8.2% in the general population, 7.3% in those exposed to cattle and 61% in pulmonary TB household contacts. After adjusting for confounding and sampling design, exposure to cattle and exposure to pulmonary TB were associated with higher risk of LTBI than the general population (respectively odds ratio (OR): 3.56, 95%CI: 0.34 to 37.03; and OR: 10.36, 95%CI: 3.13 to 34.21). Children frequently consuming cow milk had higher risk of LTBI (OR: 3.35; 95%CI 0.18 to 60.94).

**Conclusion:** Despite limited statistical power, this study suggests that children exposed to cattle in a setting endemic for bTB had higher risk of LTBI, providing indirect evidence that *Mycobacterium bovis* may contribute to TB burden.

**Author Summary:** Tuberculosis (TB) is the top infectious disease killer worldwide. *Mycobacterium bovis* (*M. bovis*) is the most common zoonotic and second most common cause of TB in humans. The pathogen is naturally resistant to pyrazinamide, a key component of standard anti-tuberculosis treatment, thus can hamper TB control and elimination efforts. *M. bovis* is endemic in cattle in this setting, but there is limited information on its contribution to TB burden. We used a specific test, the Interferon Gamma Released Assay, to compare latent TB infection (LTBI) rates in a random sample of children with high exposure to cattle, to that of children from the general community and from households with known pulmonary TB patient in and around a major urban centre in Cameroon; LTBI in children provide insight on recent infection, thus transmission. After adjusting for background differences, we found that exposure to cattle was associated with over three times higher risk of LTBI compared to the general population (and household exposure associated with over 10 times higher risk of infection). Our results suggest that exposure to cattle (therefore *M. bovis*) contributes significantly to TB burden, and should be investigated thoroughly to support control efforts.

## Introduction

*Mycobacterium bovis (M. bovis)* is the most common zoonotic, and second most common aetiologic cause of tuberculosis in humans after *Mycobacterium tuberculosis* (*M. tuberculosis) [1,2]*. Its natural resistance to pyrazinamide, a key component of TB standard treatment, and diagnostics difficulties, contribute to poorer treatment outcomes in patients and challenges to TB control programmes [3]. Cattle is the main animal reservoir, though *M. bovis* can also be found in a range of other mammals [4-6]. In sub-Saharan Africa, including Cameroon, strong evidence suggest that the burden of bovine tuberculosis (bTB) is on the rise, partly due to the intensification of cattle livestock farming to meet the increasing demands to higher rates of urbanisation. A 2016 study which sampled over 2300 bovine carcasses in abattoirs across the four main areas of livestock farming in Cameroon found a high prevalence of *M. bovis* ranging from about 3% in the North West region to over 21% in the Northern Region, as well as a non-negligible prevalence of non-tuberculous mycobacteria [7]. The study also reported a wide genetic diversity, and evidence of recent and expanding *M. bovis* transmission in intensive dairy farming areas in the North-West region [8].

Rapid urbanisation and increased demand for fresh dairy products has been driving a rapid expansion of the local dairy farming industry in many sub-Saharan African countries, contributing to the increased potential risk of zoonotic TB in the general population, beyond the occupational risk often limited to livestock workers. The World Health Organization (WHO) estimates the highest morbidity and mortality due to zoonotic TB, occurs in its African region, which is likely severely underestimated due to diagnostic limitations [1]. Unfortunately, in Cameroon, like most sub-Saharan African countries, zoonotic TB remains largely neglected [9], with control limited to regulatory inspection of animal carcases at the point of slaughter, mandated carcass destruction and recommended milk pasteurisation, often poorly enforced.

Despite high bovine TB prevalence, very little data exist on the impact on human population health, and its contribution to TB burden in Cameroon which limits advocacy efforts as well adequate planning and tailored control efforts [10]. Routine TB diagnosis platforms (microscopy, culture and Gene-Xpert) are mostly focused on pulmonary tuberculosis, and unable to differentiate *M. bovis* from *M. tuberculosis*. Besides, the natural history and long latency period between *Mycobacterium* infection (Latent Tuberculosis infection - LTBI) and the onset of active disease complicates correlating human disease burden from routine surveillance with contemporaneous animal epidemiology. Conversely, LTBI in children is a reliable indicator of recent *Mycobacterium* transmission [10]. We conducted a pilot study to compare the LTBI risk in children living in areas with high cattle density to that in children sampled from the general population, in and around Yaoundé, the capital and second-largest city of Cameroon. We hypothesized that higher *M. bovis* transmission and burden would be reflected by higher LTBI risk in children with more exposure to cattle and consumption of fresh cow-milk dairy products.

## Methods

### 1. Study design, setting, participants and Ethical consideration

#### 1.1 Study design, setting and participants

This cross-sectional study was conducted from December 2020 to February 2021. Participants were children aged 2 to 15-year-old with no history of diagnosis or treatment for tuberculosis; younger children under 2 year-old were not included due to low acceptability of blood sample collection. Participants were selected using systematic random sampling, stratified by age groups (2 to 5-year-old, 6 to 10-year-old and 11 to 15-year-old), sex, and place of residence (urban and peri-urban area), and from three TB exposure groups, respectively:

i. children with regular exposure to cattle: selected from the peri-urban area (Efoulan in Akonolinga health district) where a large population of nomadic Mbororo Fulani engages in livestock keeping;
ii. children from the general population, selected from Emana, Yaoundé (urban) and Menguem-si, Akonolinga (peri-urban);
iii. children from households with known pulmonary tuberculosis (PTB) cases; this group was selected to estimate LTBI prevalence in children with known recent and high exposure to TB. PTB patients under treatment, with eligible children in their household, were identified from Yaoundé’s principal TB treatment centre (Jamot Hospital).

#### 1.2 Ethical consideration

Written informed consent was obtained from parents or legal guardians, and verbal assent from children aged over 12-year-old and participant confidentiality was respected throughout the study. The study was approved by the ethics committee of the Human Health Research Council of the Centre Region in Cameroon (EC N01903/CRERSHC/2020). Administrative authorisations were also obtained for each study site.

## 2. Data collection

A face-to-face structured questionnaire was administered using the Open Data Kit suite (ODK) on a tablet computer, to collect information on participants’ socio-demographic characteristics including parental education and assets ownership, vaccination history, respiratory health history, exposure to cattle, consumption of fresh cow-milk dairy products, and household history of tuberculosis. Their upper arm was also examined for the BCG vaccine scar, with a brief clinical examination for signs of active tuberculosis.

We also collected from each participant 5ml of peripheral venous blood in a sodium heparin tube to diagnose latent TB infection using the Quantiferon-TB Gold Plus (QTF-plus) Interferon Gamma Released Assay (IGRA) (Qiagen Strasse, Germany). This assay measures the cell-mediated immune response elicited by infection with Mycobacterium tuberculosis complex, including *M. bovis*, using a mixture of early secretory antigenic peptide-6 (ESAT-6) and culture filtrate protein-10(CFP-10) antigens; this antigen mixture allows the assay to distinguish infected individuals from those uninfected, with and without Bacille Calmette and Guerin( BCG)-vaccination [11]. Stool samples were also collected to screen intestinal helminth parasites, as there has been suggestions that they can interfere with the IGRA response [12].

The blood samples were kept in the field in an ice box (0 to 4°C) and transported to the laboratory on the same day. For each sample, 1ml of blood was immediately transferred into each of four QuantiFERON-TB Gold Plus tubes (Nil tube, TB1 tube, TB2 tube, and Mitogen tube) and incubated for 24 hours at 37°C temperature. The tubes were then centrifuged at 3000G for 15 minutes, the plasma was collected, aliquoted and frozen at -20°C until analysis. At the end of field data collection, the stimulated plasma samples were analysed using quantitative Enzyme Linked Immuno-Assays (ELISA) according to the manufacturer’s instructions. The manufacturer’s software was then used to interpret the raw optical densities data, including the assay validation and quality control, and computation of interferon gamma (IFN-ϒ) concentration in international units per millilitre (IU/ml). A positive IGRA result was defined as valid assay with IFN-ϒ released concentration ≥ 0.35 IU/ml after blood stimulation with mycobacterial antigens.

## 3. Statistical analysis

The primary outcome, LTBI, was defined as a positive QFT-plus result with no clinical sign of active tuberculosis. The main exposure was categorised into three TB groups: exposure to a known TB case through household membership, frequent exposure to livestock, with children from the general community being the comparison baseline. We also performed exploratory analyses for the association between various participant characteristics and LTBI.

The questionnaire data was cleaned and coded, then consistency and range-checks performed to check the data quality and correct data-entry mistakes. The distribution of categorical variables was examined, with some categories merged as required to the small sample size. We performed descriptive analyses using simple cross-tabulations, looking at the distribution of various characteristics in the study participants. Principal Component Analysis (PCA) was used to combine information on assets ownership into a socio-economic score [13]; assets used in the score included ownership of a television, cable TV, computer, smartphone, fridge, gas stove, and the house building material and number of bedrooms. The PCA predicted score was categorised into lower versus higher socio-economic status.

We calculated the crude LBTI prevalence (without adjusting for sampling stratifying variable age, sex, and place of residence), tabulated against participant’s characteristics, and we investigated its association with the TB exposure group and other participants’ characteristics. The magnitude of association was measured using Odds Ratios (ORs) and 95% confidence intervals (95% CI). First, we computed minimally adjusted ORs (and 95% CI) of association between LTBI and variables, controlling for age, sex and place of residence. We then built a fuller multivariable logistic regression model, further adjusting for confounding by other participant characteristics. The model was constructed using a forward stepwise approach, starting with the minimally adjusted model (including TB exposure group, age group, sex and place of residence as a priori confounders), then adding other variables in turn and checking their introduction did not cause data sparsity or collinearity.

We also did a subgroup analysis excluding children with exposure to TB patients in their household, thus restricted to those exposed to livestock and from the general community, to investigate the association between consumption of fresh cow-milk dairy product and raw meat and LTBI.

All statistical tests of association were done using the likelihood ratio test (LRT), and OR and 95% CI were obtained using maximum likelihood estimation. Statistical analyses were conducted using Stata 17.

## Results

### 1. Participant’s characteristics

We recruited 160 children, respectively 41 (26%) with frequent exposure to livestock, 46 (29%) from households with a TB case, and 73 (45%) from the general community. 89 (56%) study participants were female, and 68 (42%) were aged 2 to 5-year-old. About half the participants lived in the urban area, and 50% were BCG vaccinated, although vaccine uptake was noticeably lower in children exposed to livestock (2.4% versus 67.1% in general community children and 63.0% in household contacts). Only 7 (4.4%) children had evidence of intestinal helminths in their stool. The consumption of dairy products from fresh cow milk was more common in children exposed to livestock (98%). The detailed characteristics of participants are presented in Table 1.

**Table 1.** Characteristics of study participants overall and by exposure group.

| Variables | Overall (%) | General community (%) | Exposed Livestock (%) | Household exposure (%) |
| --- | --- | --- | --- | --- |
| **Age group (Years)** |  |  |  |  |
| **[2-5]** | 68 (42.5) | 29 (39.7) | 19 (46.3) | 20 (43.5) |
| **[6-10]** | 54 (33.7) | 26 (35.6) | 14 (34.1) | 14 (30.4) |
| **[11-15]** | 38 (23.7) | 18 (24.7) | 8 (19.5) | 12 (26.1) |
| **Sex** |  |  |  |  |
| **Female** | 89 (55.6) | 40 (54.8) | 24 (58.5) | 25 (54.3) |
| **Male** | 71 (44.4) | 33 (45.2) | 17 (41.5) | 21 (45.6) |
| **Residence area** |  |  |  |  |
| **Peri-urban** | 81 (50.6) | 40 (54.8) | 41 (100) | 0 (0) |
| **Urban** | 79 (49.4) | 33 (45.2) | 0 (0) | 46 (100) |
| **Socio-economic level** |  |  |  |  |
| **Low** | 82 (51.2) | 38 (52.0) | 41 (100) | 3 (6.5) |
| **High** | 78 (48.7) | 35 (47.9) | 0 (0) | 43 (93.5) |
| **Head of household education level** |  |  |  |  |
| **Primary** | 82 (51.2) | 29 (39.7) | 41 (100) | 12 (26.1) |
| **Secondary** | 78 (48.7) | 44 (60.3) | 0 (0) | 34 (73.9) |
| **BCG vaccination status** |  |  |  |  |
| **No** | 81 (50.6) | 24 (32.9) | 41 (97.6) | 17 (37) |
| **Yes** | 79 (49.4) | 49 (67.1) | 1 (2.4) | 29 (63.0) |
| **Household cattle ownership** |  |  |  |  |
| **No** | 11 (73.7) | 73 (100) | 0 (0) | 45 (97.8) |
| **Yes** | 42 (26.3) | 0 (0) | 41 (100) | 1 (2.2) |
| **Stool parasites results** |  |  |  |  |
| **Negative** | 153 (9.2) | 67 (91.8) | 41 (100) | 45 (97.8) |
| **Positive** | 7 (4.4) | 6 (8.2) | 0 (0) | 1 (2.2) |
| **Consumption of boiled cow milk** |  |  |  |  |
| **No** | 117 (73.1) | 72 (8.6) | 1 (2.4) | 44 (95.6) |
| **Yes** | 43 (26.9) | 1 (1.4) | 40 (97.6) | 2 (4.3) |
| **Consumption of crude cow milk** |  |  |  |  |
| **No** | 154 (96.2) | 72 (98.6) | 37 (90.2) | 45 (97.8) |
| **Yes** | 6 (3.7) | 1 (1.4) | 4 (9.8) | 1 (2.2) |

### 2. Latent tuberculosis infection prevalence and risk factors

Overall, the prevalence of latent tuberculosis infection in our study participants was 23.1% (37/160). Before adjusting for the sampling stratifying variables age, sex and place of residence, the crude prevalence was respectively 60.9% (28/46) in children exposed to a TB patient in their household, 7.3% (3/41) in children exposed to livestock and 8.2% (6/73) in children randomly selected from the community. The prevalence was higher in male (25.4%) than female (21.4%), and about twice as high in older 11-15 years old children (36.8%) than younger 6-10 years (18.5%) and 2-5 years (19.1%) children. We also found a higher prevalence in children from urban areas (41.8%) than peri-urban areas (4.9%). The crude LTBI prevalence by study characteristics is presented in Table 2.

**Table 2.** Crude prevalence of latent tuberculosis infection by participants’ characteristics.

| Variables | N | IGRA positive (%) |
| --- | --- | --- |
| **Exposure group** |  |  |
| General community | 73 | 6 (8.22%) |
| Exposed to Livestock | 41 | 3 (7.32%) |
| Household exposure | 46 | 28 (60.87%) |
| **Age group (years)** |  |  |
| [2-5 ] | 68 | 13 (19.12%) |
| [6-10 ] | 54 | 10 (18.52%) |
| [11-15 ] | 38 | 14 (36.84%) |
| **Sex** |  |  |
| Female | 89 | 19 (21.35%) |
| Male | 71 | 18 (25.35%) |
| **Residence area** |  |  |
| Peri-urban | 81 | 4 (4.94%) |
| Urban | 79 | 33 (41.77%) |
| **Socio-economic level** |  |  |
| Low | 82 | 6 (7.32%) |
| High | 78 | 31 (39.74%) |
| **Head of household education level** |  |  |
| Primary | 82 | 11 (13.41%) |
| Secondary | 78 | 26 (33.33%) |
| **BCG vaccination status** |  |  |
| No | 81 | 18 (22.22%) |
| Yes | 79 | 19 (24.05%) |
| **Consumption of boiled cow milk** |  |  |
| No | 117 | 33 (28.21%) |
| Yes | 43 | 4 (9.30%) |

After adjusting for age, sex and place of residence, there was evidence of association between the exposure group and LTBI (p < 0.001), with higher risk of LTBI in children exposed to livestock (OR: 3.56, 95% CI: 0.34 to 37.03) and those exposed to TB patient in their household (OR: 10.36, 95% CI: 3.13 to 34.21), compared to children from the general community. These associations remained mostly unchanged after further adjustment for socio-economic level, head of household education level and BCG vaccination status. The odds of LTBI also appeared higher in children from urban area than peri-urban area (fully adjusted OR: 7.39, 95% CI: 0.58 to 94.73, p = 0.092), and increased with age (fully adjusted OR: 1.06, 95% CI: 0.32 to 3.47; and OR: 3.59, 95% CI: 1.03 to 12.60), respectively in 6-10 years and 11-15 years compared to 2-5 years, and p-value for trend =0.056. BCG vaccination was associated with lower risk of LTBI, albeit with wide confidence interval (fully adjusted OR: 0.53, 95% CI 0.17 to 1.65). Full results are in Table 3.

**Table 3.** Association between participants’ characteristics and latent tuberculosis infection.

| Variable | Crude OR | 95% CI | p-value | Adjusted OR | 95% CI | p-value |
| --- | --- | --- | --- | --- | --- | --- |
| **Exposure group** |  |  |  |  |  |  |
| **General community** | 1 |  |  | 1 |  |  |
| **Exposed to Livestock** | 3.56 | 0.34 - 37.03 | < 0.001 | 3.59 | 0.28 - 45.52 | < 0.001 |
| **Household exposure** | 10.36 | 3.13 - 34.21 |  | 10.11 | 2.98 - 34.35 |  |
| **Age group (years)** |  |  |  |  |  |  |
| **[2-5]** |  |  |  | 1 |  |  |
| **[6-10]** | 0.88 | 0.32 - 2.43 | 0.111 | 1.06 | 0.32 - 3.47 | 0.056 |
| **[11-15]** | 2.44 | 0.87 - 6.79 |  | 3.59 | 1.03 - 12.60 |  |
| **Sex** |  |  |  |  |  |  |
| **Female** | 1 |  |  | 1 |  |  |
| **Male** | 1.36 | 0.58 - 3.19 | 0.474 | 1.22 | 0.45 - 3.26 | 0.686 |
| **Residence area** |  |  |  |  |  |  |
| **Peri-urban** | 1 |  |  | 1 |  |  |
| **Urban** | 14.18 | 4.65 - 43.22 | < 0.001 | 7.39 | 0.58 - 94.73 | 0.092 |
| **Socio-economic level** |  |  |  |  |  |  |
| **Low** | 1 |  |  | 1 |  |  |
| **High** | 1.41 | 0.34 - 5.88 | 0.633 | 0.95 | 0.15 - 5.87 | 0.948 |
| **Head of household education level** |  |  |  |  |  |  |
| **Primary** | 1 |  |  | 1 |  |  |
| **Secondary** | 1.35 | 0.51 - 3.58 | 0.545 | 1.90 | 0.54 - 6.65 | 0.312 |
| **BCG vaccination status** |  |  |  |  |  |  |
| **No** | 1 |  |  | 1 |  |  |
| **Yes** | 0.41 | 0.16 - 1.08 | 0.068 | 0.53 | 0.17 - 1.65 | 0.274 |

In the subgroup analysis excluding children exposed to TB patients in their household, we found higher odds of LTBI in children who reported frequent consumption of boiled cow milk, though numbers were small, and the confidence interval was wide (fully adjusted OR: 3.35; 95% CI :0.18 to 60.94) ( Table 4.).

**Table 4.** Association between consumption of fresh cow-milk products, raw meat, and latent tuberculosis infection in children exposed to livestock and from the general community.

| Variable | LTBI prevalence (%) | Crude OR | 95% CI | p-value | Ajusted OR | 95% CI | p-value |
| --- | --- | --- | --- | --- | --- | --- | --- |
| **Boiled milk** |  |  |  |  |  |  |  |
| **No (n=73)** | 6 (8.22%) | 1 |  |  | 1 |  |  |
| **Yes (n=41)** | 3 (7.32%) | 2.07 | 0.29 - 14.52 | 0.462 | 3.35 | 0.18 - 60.94 | 0.413 |
| **Yoghurt** |  |  |  |  |  |  |  |
| **No (n=102)** | 8 (7.84%) | 1 |  |  | 1 |  |  |
| **Yes (n=12)** | 1 (8.33%) | 0.94 | 0.15 - 5.67 | 0.943 | 0.89 | 0.13 - 5.94 | 0.905 |
| **Raw meat** |  |  |  |  |  |  |  |
| **No(n=80)** | 7 (8.75%) | 1 |  |  | 1 |  |  |
| **Yes (n=34)** | 2 (5.88%) | 0.48 | 0.05 - 4.82 | 0.535 | 0.32 | 0.02 - 4.68 | 0.406 |

## Discussion

This pilot study contributes some insights in the potential contribution of bovine tuberculosis, a neglected zoonosis, to TB burden in Cameroon, especially in children. Key findings were suggestions that after adjusting for confounding, children with frequent exposure to cattle may have over three-time higher risk of LTBI than those from the general community. Despite our study’s modest power, these results, in a context where Bovine TB( bTB) is endemic and highly prevalent in cattle, are consistent with the hypothesis of spill over to human populations, thus contributing to the TB burden. There is limited data on the true burden of bTB in human populations in Africa, but the results are also aligned with reports from other sub-Saharan African settings where higher TB rates have been observed in pastoral populations or with occupational exposure, 21% compared to the general population[14]. The LTBI prevalence in children exposed to cattle husbandry in this study is however lower than what has been reported in people with occupational risk: Torres and colleagues in Mexico reported 58.5% of individuals with latent TB infection [15]; Catalina and associates reported in a group of 674 agricultural workers exposed to livestock in Columbia 10.7% [16]. The difference likely reflect different populations and exposure, with the later groups mostly made of adults working closely and frequently with cattle, thus more exposed.

Subgroup analyses showed that the consumption of cow’s milk was associated with higher risk of infection in our study population. This is a common practice in pastoralist communities [17,18], and a cheap and readily available source of nutrients for developing children. It suggests that exposure to contaminated cow food products, especially milk, could be an important bTB transmission route in our target population. If confirmed, this finding offers practical targets for risk mitigating public health actions.

An incidental finding in our pilot study is the very high (61%) LTBI prevalence in children exposed to PTB cases in their households. There is limited data measuring LTBI using IGRA in children household contacts. Data from a high TB burden province in South Africa, using the Tuberculin Skin Test (TST) which is less specific, reported lower LTBI prevalence of around 20% in children under 15 years [19]. The higher prevalence in our study could reflect delays in pulmonary TB patients seeking care, or challenges in the implementation of national TB policies regarding the screening and prophylactic treatment of household contacts; this finding should be explored further and may offer additional insights into factors contributing to persisting high TB rates in urban settings despite control efforts.

To the best of our knowledge, this study is the first to investigate LTBI among children in Cameroon using IGRA. Participants were recruited during the Covid-19 pandemic when restrictions were in place to minimise social mixing; this presented challenges with response rate and implementation, partly explaining the small sample size. There was however no suggestion that non-response was different between the targeted subgroups, therefore mostly leading to study low statistical power, rather than biased results. The small study size also limited or ability to explore the association with levels of exposure in more details. Using IGRA, especially the QFT-plus assay offered higher specificity in the identification of LTBI patients compared to studies using TST, in a population with high BCG vaccination coverage, and we also checked for intestinal helminths to account for any interference with assays. We were also able to adjust analysis for several confounders, including BCG vaccination and socio-economic level. It is possible that there was residual confounding, given we only used approximate measures such as the BCG scar rather than vaccination records, and assets ownership; but their confounding effect appeared modest, so it is unlikely residual confounding would have biased the results. In summary, while we appreciate the limitations of this pilot study, notably its limited statistical precision, we believe it does provide invaluable data on an understudied and neglected, yet important public health issue.

## Conclusion

Overall, by showing higher LTBI risk in children exposed to cattle than the from general population, our study provides indirect evidence in support of the hypothesis that bovine tuberculosis may contribute to the TB burden in Cameroon. This should be explored in a larger study, including a follow-up component to monitor progression to active tuberculosis which could allow the confirmation of *M. bovis* as the etiologic agent and an estimation of its net contribution to disease burden. We would also advise the Cameroon National TB control programme to further investigate the seemingly high LTBI prevalence in children household contacts of people with pulmonary tuberculosis, as part of its efforts to reduce TB transmission.

## Data Availability

The data that support the findings of this study will be publicly available from the London School of Hygiene and Tropical Medicine public data repository accessible at https://datacompass.lshtm.ac.uk/

## Acknowledgments

We would like to acknowledge and thank all study participants and their families, without whose generous cooperation this study would not be possible. We also thank the head of the Laboratory of Tuberculosis Research Kolbisson (Professor Penlap Beng Véronique) manager of the Wise project Cameroon. We also thank the Directors of the Jamot hospital in Yaounde, the Akonolinga, Djoungolo and Nkoldongo health districts. We thank all the colleagues and personnel who have contributed to this work, including, Djune Yemeli Linda for her assistance with her materials; Mbickmen Tchana Steve, Balog Aubin, Bamou Heumou Louis Rolf, Mbakam Laetitia for their laboratory support; Achobi Alain and Youssoufa for their always presence in the field.

## Authors’ contributions

PND conceives the study; MAFT and PND designed the study protocol, with input from HCND, JK, JFTK and HDM. MAFT, HOTP, RD and MON performed field data collection under supervision from PND; MAFT, MAN and PND conducted statistical analyses. MAFT and PND prepared the manuscript, with contribution from all co-authors.

